# Exploring Barriers to Parent-Adolescent Sexual-Risk Communication among Adolescents in Port Harcourt Nigeria: Adolescents’ and Parents’ Perspective

**DOI:** 10.1101/2024.04.13.24305352

**Authors:** Okpalaku Joycelyn Chidinma, Chigozirim Ogubuike

## Abstract

**Background:** Adolescent risky sexual behaviour is a public health problem with its deleterious outcomes. Parents are the most influential source of sexuality education to adolescent, yet adolescents lack sexuality education. The study explored barriers in parent-adolescent sexual-risk communication from both perspectives in Port-Harcourt LGA, Rivers State, Nigeria.

**Materials and Method:** A cross-sectional study design using explanatory sequential mixed methods approach was implemented. Three hundred and twenty nine in-school adolescents participated in the quantitative study and recruited using multi-stage sampling technique while 9 parents of adolescents and 16 adolescents participated in the qualitative study. A semi-structured administered questionnaire was used to elicit information from the adolescents while an FGD and IDI guide was used to elicit information from in-school adolescents and parents respectively. The qualitative data was analysed using descriptive statistics and chi-square while the qualitative data was subjected to thematic analysis.

**Results:** The mean age of the adolescents was 16.0±1.1 years and 55% were males. A quarter (21%) of the parents had never discussed sex with their adolescents. The barriers identified from the adolescents’ perspective were parental factors (parents being too busy, judgmental, low knowledge), individual factors (discomfort to initiate communication, lack of trust), religious and cultural factors. The barriers from the parents’ perspective were shame to initiate communication, fear of outcome, feeling children are too young and lack of accurate information.

**Conclusion:** The barriers to parent-adolescent communication featured interplay of parental, individual, cultural and religious factors. Parents should be trained to initiate timely and accurate sexuality education to the adolescent to curb adolescent risky sexual exploitations.

## BACKGROUND

Adolescence, which occurs between the ages of 10 and 19 years, is a stage between childhood and adulthood marked by profound psychosocial, psychological, and social changes.^1, 2^ Adolescents constitute of 1.8 billion or one-fourth of the world’s population, therefore the care for this group is accompanied with socioeconomic progress and societal advancement that are being confronted. ^3^ They go through anatomical and psychological changes that have a profound impact on their mental wellness, which is manifested as high levels of sexual curiosity. ^4^ This sexual desire could result in risk-taking sexual behaviour if there is no behaviour counselling during the adolescent years. ^5^ Risky sexual behaviours are behaviours that put a person at risk for STIs like HIV and unintended pregnancies. These actions include having several sexual partners, transactional sex, forced or coerced sex, and unprotected sexual contact. ^6^ Adolescents who are sexually active are more likely to participate in risky behaviours that could raise the likelihood of early sexual activity, increase the chance of unintended pregnancy and abortion, and increase the risk of sexually transmitted diseases. ^7^ Additionally, they are vulnerable to several issues and attitudes related to sexual and reproductive health. ^8^

Research has shown that open conversation between parents and children about sex can significantly lower the sexual risk that adolescents face. ^9, 10^ Adequate sexuality communication between young people and their parents or other caregivers has been found to be one of the elements that protects against non-risky sexual behaviours, such as a delayed sexual debut, especially in females. ^11^ Open communication between parents and adolescents has been proven essential for lowering the morbidity and mortality associated with risky sexual behaviours. ^12^ Also, parent-child discussion on sexual and reproductive topics, have shown to increase awareness and reduce risk-taking behaviour. ^13^ According to the Advocates for Youth, parent-adolescent sexual-risk communication about contraception, the menstrual cycle, premarital sex, unintended pregnancies, and ways to prevent STDs helps solve problems associated to sexual and reproductive health. ^14^ Nevertheless, it is reported that the majority of the knowledge that adolescents have about sex or sex education comes from their female classmates and schoolmates, who rarely provide accurate or precise information. ^15^ Research has shown that the sexual behaviour of adolescents is always a sensitive issue in churches, schools, and families with each of them pointing fingers at the other when problems that relate with adolescent sexual behaviour crop up. ^16^

Most parents find it difficult to discuss sexual and reproductive health issues with their children and instead choose to steer the conversation toward “safe” topics or provide cautionary statements whenever there is a sexually related issue. ^17^ Parents and adolescents’ communication or lack of it has always arisen to serious social and health consequences which include parent child conflict, school dropouts, unwanted pregnancy, unsafe abortion, STIs such as HIV and AIDS. Most parents experience discomfort when speaking about adolescent reproductive health with their adolescents and this can hinder the occurrence of effective reproductive health communication.^11^ Parent’s generally have a negative aversion to communication on sex education and this aversion to the teaching of sexual health by parents is due to a strong proclivity to culture and religious teachings imbibed from their fore parents. ^18^ Parental and societal distaste to sex education hasn’t changed over time which has resulted into an overlooking of adolescents as they receive virtually no or relatively little reproductive health care and education. ^19^ This study explored parent-adolescent sexual risk communication from both the adolescent and adolescent perspective to unravel the barriers; this would help in policy and programmatic directions curb risky sexual exploitations in adolescents.

This study focused on parent-adolescent communication on sexual risk practices because it is the researcher’s conviction that adolescents are greatly influenced by their parents or caregivers which are the most important source on sex education and prevails over other factors that pertain to adolescents. Therefore, this study was conducted to determine the proportion of adolescents that received sexuality education from their parents, explore barriers to parent-sexual risk communication and recommendations for easier sexuality communications by parents.

## MATERIALS AND METHOD

### Study Area

Port-Harcourt city is the capital of Rivers State in Nigeria otherwise known as Port Harcourt Local Government Area (PHALGA). It is one of the 23 local government areas created for the state. It is located in the southern part of the country and is one of the states in the Niger Delta region. The Local Government Area is bounded by Ikwerre and Etche Local Government Areas at the North, Asari-Toru and Okrika Local Government areas at the South, Emohua LGA at the West and Eleme and Oyigbo LGAs at the East in Rivers State. ^20^ In 2021, the population of Port Harcourt was estimated to be 3,171,076. ^21^ Port Harcourt is the nerve centre of the Nigerian Oil industry and includes other industries too. It is also the nation’s second largest sea port with the Onne Port Complex in close proximity. Marine agriculture is the main occupation of the people of Rivers state with farming and trading being the two major occupations of Port Harcourt residents. The state has a 154 non-tertiary public, private and government schools which means a whole lot of adolescents enrolled into these schools which covers the study population for this study.

### Study Design and Population

A cross-sectional study design using the explanatory sequential mixed method approach was utilized for this research. It is a two-phase mixed methods design that starts with the collection and analysis of quantitative data followed by the subsequent collection and analysis of qualitative data. The study population involved in-school adolescents and parents of adolescents. This study included in-school male and female adolescents between the ages of 15-19 years from junior secondary school (JSS3) and senior secondary school (SS1, SS2 and SS3) in selected public and private schools in Port Harcourt LGA. Parents recruited were those that have adolescents. The adolescents were involved in both quantitative and qualitative data collection while the parents were involved in only qualitative data collection.

### Sample size determination

The sample size for the quantitative aspect was determined using Cochran’s formula:

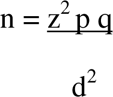

Where, n =the desired sample size.

z = the standard normal deviate, usually set at 1.96 which corresponds to 95% confidence level. p = the proportion of adolescents who communicated with their parents on SRH issues was 36.9 or 37% (Shiferaw et al.) ^15^

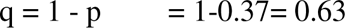

d = degree of accuracy desired, usually set at 0.05.

n= 1.96^2^ x 0.37 (1-0.37)/0.05^2^

= 358. 1. Assuming a non-respondent rate of 10%, the minimum sample size was estimated at 394.

For the qualitative aspect, 9 parents of adolescents and 16 adolescents were purposively recruited to participate in the study.

### Sampling method

For the quantitative aspect a multistage sampling method was used in this study. The first stage involved random selection of schools. The list of schools in Port Harcourt LGA was obtained from the Rivers State Ministry of Education and used as the sampling frame. The schools were first stratified into private and public schools and simple balloting was used to select two private and two public schools. The second stage is the selection of students according to classes and gender. The students were stratified into 3 groups based on their classes i.e. SSS1, SSS2, and SSS3 and then re-stratified based on their sexes (Males and Females). The third stage involved the proportionate allocation. The sub-sample for each stratum was divided proportionately into the number of arms/forms in each class. The sample size was proportionately divided into each of the strata based on their population. About 100 students were recruited from each school, hence a total of 400 participants were recruited.

In the qualitative aspect the respondents were recruited purposively, the adolescents participated in a focus group discussion (FGD) while the parents participated in an in-depth interview (IDI).

### Data Collection

Recruitment for participants for the pilot study commenced on the 5^th^-7^th^ May 2021 while recruitment of participants for the study was between 6th– 24th^th^ July 2021. Parents were recruited 6^th^ −11^th^ August 2021.

Structured interviewer administered questionnaires were given to the adolescents which they completed themselves. Data was gotten from the parents via in-depth interview and was audio recorded. The consent of the participants was sought before the research questionnaire was given. Participants were asked to sign on a well-written consent form enclosed on the questionnaire after the study had been explained to them so as to show their voluntary participation. For adolescents below 18 years, it was ensured that their guardians in form of the school principals were present, signed the consent forms and the adolescents also assented before questionnaires were administered. The questionnaires were collected after being completed and the participants were appreciated for their contribution and time.

### Study instrument and Data analysis

A pre-tested structured interviewer-assisted questionnaire was used to elicit information on the socio-demographic characteristics of the respondents, source of information and proportion of adolescents that received sex education. The researcher administered the questions to the adolescents and retrieved them after completion. An in-depth interview guide and focus group discussion guide was used to elicit qualitative information on the barriers and facilitators to parent-adolescent sexual-risk communication. The questionnaire was adapted from earlier studies on parent-adolescent communication about sexuality, ^17, 22, 23, 24^ while some other questions were introduced by the researcher and given to two experts to check for face validity. SPSS statistical software version 25 was used for the qualitative analysis. Mean and standard deviation were used for descriptive statistics. Atlast ti software was used for the analysis of the qualitative data. The interviews were audio recorded, transcribed verbatim, carefully read and edited by the researchers to ensure original meanings were not lost to translation. The qualitative data was subjected to thematic analysis.

### Ethical considerations

Ethical clearance was obtained from the Research and Ethics Committee of the University of Port Harcourt and permission obtained from the school authority of chosen schools before commencement of the study. A written consent was also obtained from the parents/guardians before starting the study. Assent was gotten for the students below 18 years of age in addition to the written consent signed by their guardians (School Principal) while adolescents 18 years and above completed the consent forms themselves. Information of the study was made available on each consent form. Confidentiality was guaranteed as personal details of respondents were not included in the data collection instrument. Serial numbers were used and as a result, collected data wasn’t linked to any of the respondents.

## RESULTS

### Socio-demographic characteristics

#### Socio-demographic characteristics of the adolescents

The socio-demographics of adolescents recruited into the study is shown in Table 1. A total of 329 out of 400 students completed the questionnaire giving a response rate of 83.5%. The result revealed that 236 (71.7%) of the adolescents were between the ages of 15-16 years with a mean age of 16.0±1.1 years, 190 (57.8%) were males, 165 (50.2%) were in SS2 and 316 (96.0%) of the respondents were Christians. Most of the adolescents 223(67.8%) belonged to a monogamous family and 202(61.4%) lived with their biological parents. About 196(59.6%) of the adolescents had a family size of 4-6 persons, 129(39.2%) of the adolescents’ fathers had only vocational training, 102(31.0%) of the adolescents mothers had no formal education, 145(44.1%) of the adolescents’ fathers were self-employed and 204(62.0%) of the adolescents’ mothers were self-employed. For the qualitative data, 16 adolescents participated in an FGD of two groups (8 males and 8 females).

**Table 1.**
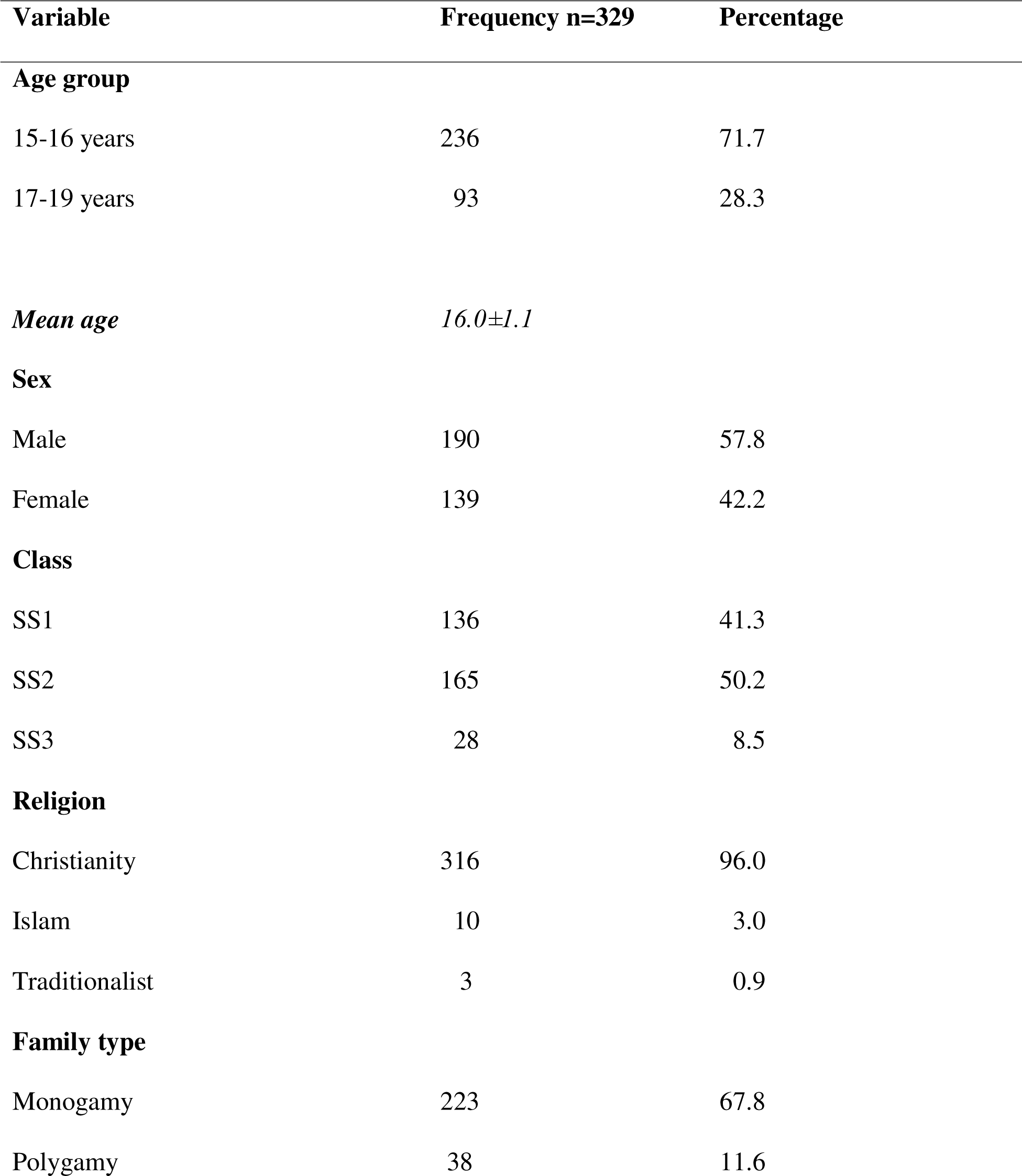
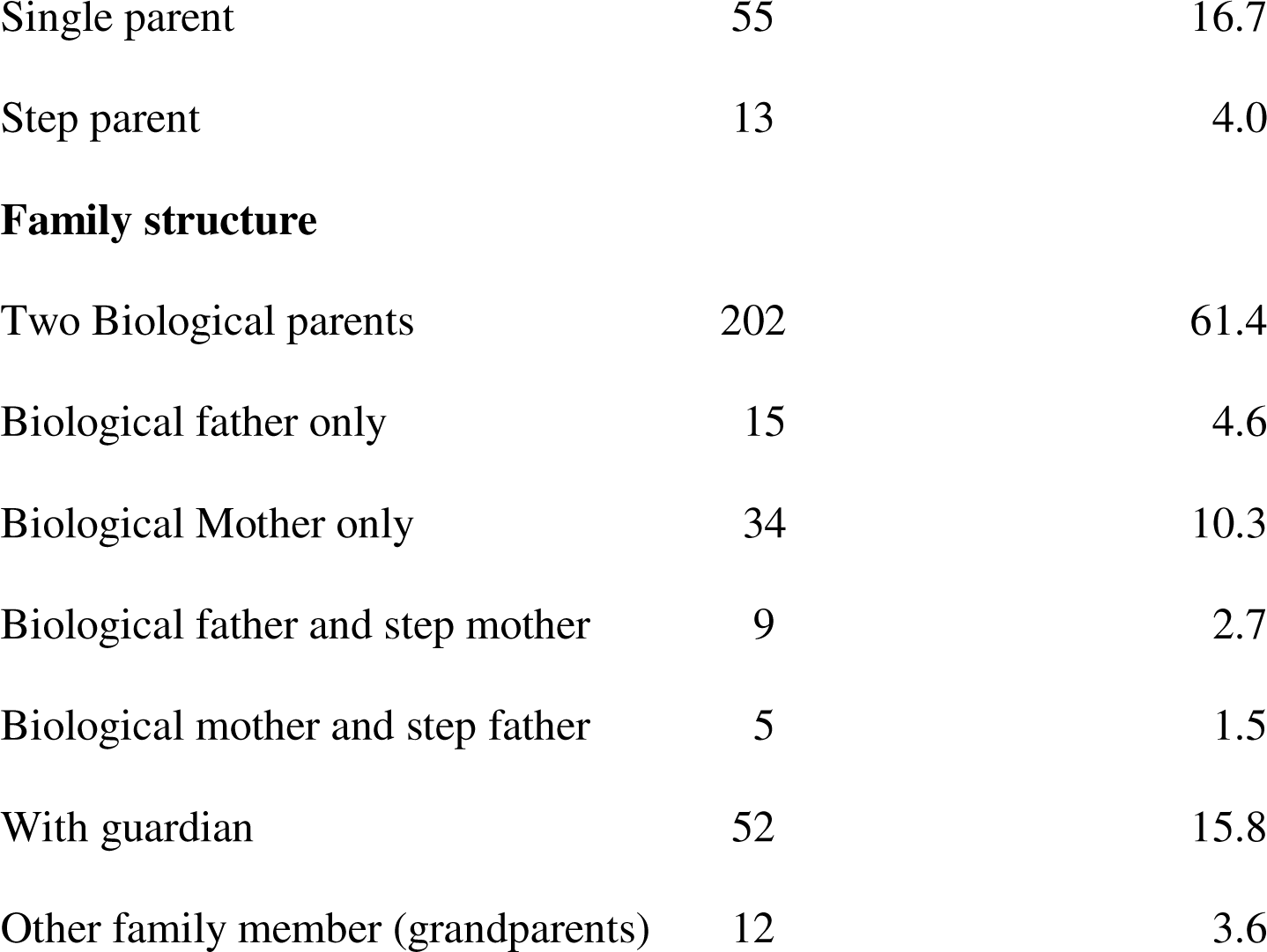
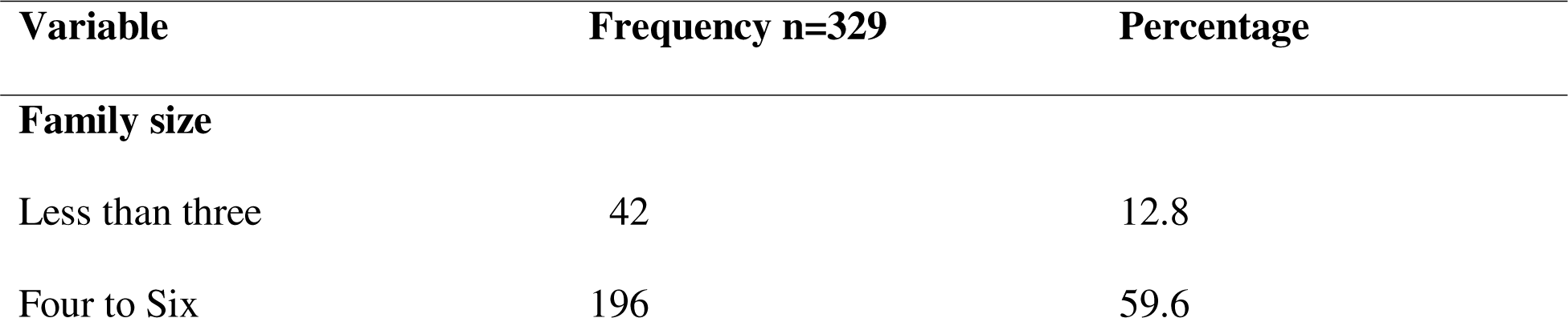
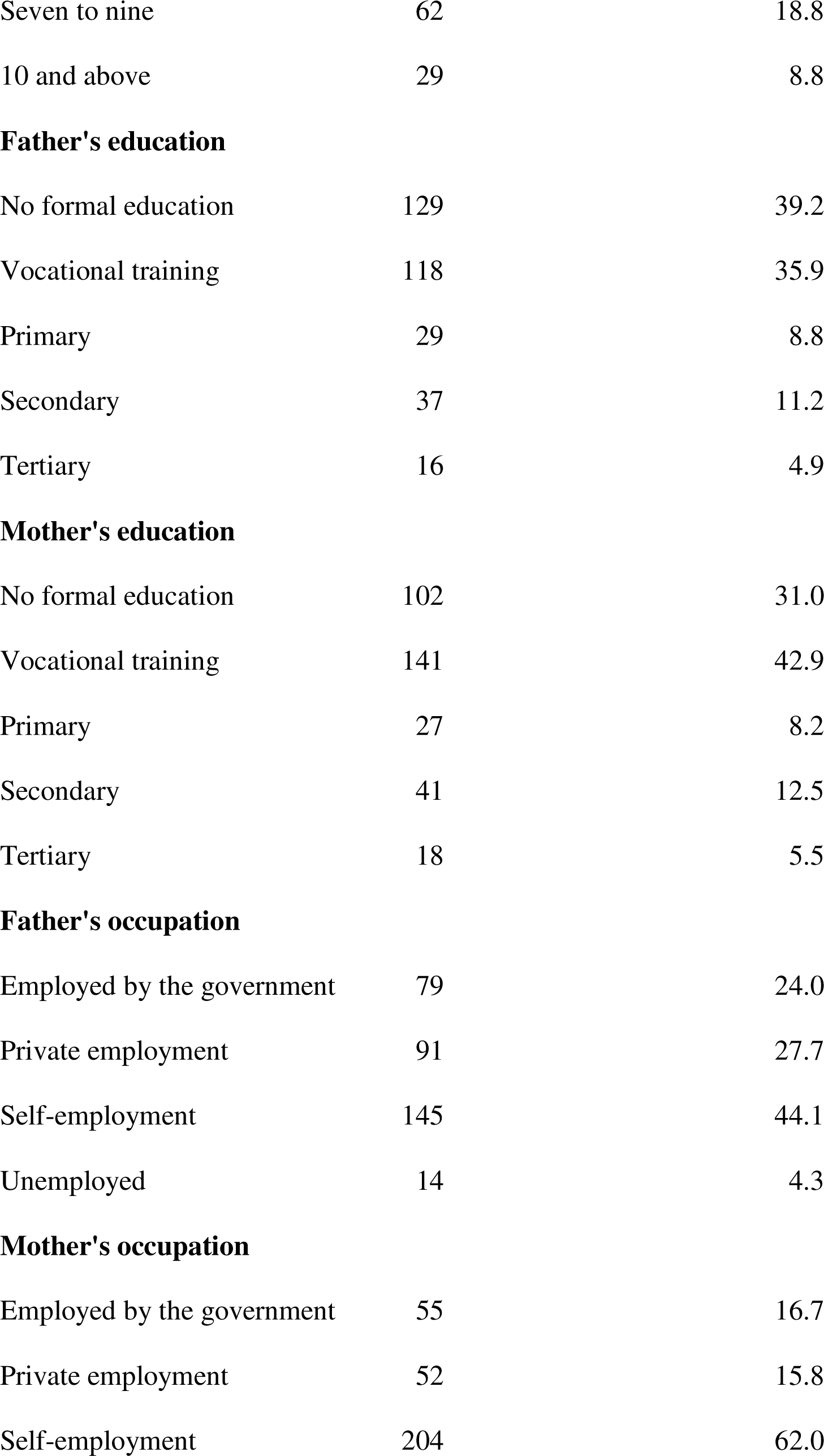
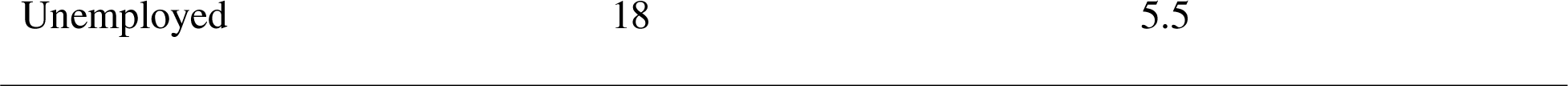
a: Socio-Demographic Characteristics of Adolescents in Port Harcourt, Rivers State b: Social Demographic Characteristic of Adolescents in Port Harcourt, Rivers State

#### Socio-demographic characteristics of parents

Nine parents of adolescents participated in an IDI session comprising of 5 males and 4 females among which 3 were teachers, 3 traders and 3 civil servants. Six of them had attained a tertiary education and three had attained a secondary education.

#### Proportion that received sexual education from parents

Table 2 shows the proportion of adolescents that received sexual education from their parents. It was shown that 69(21.0%) of the parents had never discussed sex education with their children, 107(32.5%) of adolescent reported that they would be more comfortable discussing sexual related issues with their mother. A majority of them 190(57.8%) received sexual education from school.

**Table 2:** Proportion of Adolescents in Port Harcourt, Rivers State That Received Sexual Education from Parents/guardians.

| Variable | Frequency n=329 | Percentage |
| --- | --- | --- |
| How often parents/guardian discuss sex education |  |  |
| Never | 69 | 21.0 |
| Rarely | 91 | 27.7 |
| Sometimes | 123 | 37.4 |
| Often | 20 | 6.1 |
| Always | 26 | 7.9 |
| **Comfortable discussing sexual related issue with** |  |  |
| Father | 35 | 10.6 |
| Mother | 107 | 32.5 |
| Siblings | 31 | 9.4 |
| Teachers | 17 | 5.2 |
| Peers of the same sex | 50 | 15.2 |
| Friends | 89 | 27.1 |
| **Source of sex education** |  |  |
| Home | 105 | 31.9 |
| School | 190 | 57.8 |
| Internet/social media | 25 | 7.6 |
| Church | 9 | 2.7 |

#### Perceived Barriers to Parent-adolescent Sexual-risk Communication: Adolescents’ Perspective

Adolescents outlined some of the perceived barriers to effective sexual-risk communication. The barriers were grouped into three broad factors; parental factors (parents being too busy, parents are too judgmental, parental poor/inaccurate knowledge on sex education, parental belief that discussion will initiate sex); individual factors (lack of trust, fear/discomfort with discussion) and religious/cultural factors.

##### A. Parental factors

###### Parents too busy with work

One of the barriers to effective sexual risk communication between parents and adolescents were parents being too busy with work and rarely had time to communicate with their wards.

A male adolescent had asserted that:

> My dad is hardly ever home and even when he is at home; he says he’s too tired to have any conversations with me. He doesn’t have out time. All of my sexual knowledge is either from school, my friends or from the internet.

###### Male Adolescent (16-20 years)

A female adolescent had this to say:

> My mom is always travelling for business so, discussing sex with her is almost impossible. I can’t discuss with my dad because he is of the opposite sex. Most of the knowledge I have on sex is what I was taught in school by my teachers, some form my friends too.

###### Female Adolescent (11-15 years)

Due to parents being too busy at work, adolescents’ rarely received sexual-risk communication from their parents. They mentioned receiving sexual education from the school, their teachers, peers and the internet.

###### Parents are too judgmental

All female adolescents agreed that their parents are too judgmental which is why they don’t discuss sexual and reproductive health issues with them, also their male counterpart asserted same. One of the female adolescent reported that:

> I don’t discuss sexual matters with my mom because she will just assume that I want to start having sex that I why I am asking so, I don’t bother meeting her to start up such discussions. She judges a lot and condemns me.

###### Female Adolescent (16-20) years

A male adolescent averred that he wouldn’t discuss sex with his parents because:

> My first knowledge about SRH was from school. My parents are the kind of people that read meaning into everything; even mere questions. I asking questions about these issues will mean that I have already started having sex or I want to have sex. There was this time i asked my dad if condoms were 100% effective in preventing pregnancy. My dad called my mom immediately and they started asking me all sorts of really embarrassing questions. Since then, i never asked them such questions again.

###### Male Adolescent (16-20 years)

The adolescents reported that their parents were too judgemental, read meaning into little things and condemn them when they asked questions on sexual and reproductive health related matters. They resorted never to have such discussions again with their parents.

###### Parental Poor/inaccurate knowledge on SRH issues

Some of the adolescents admitted that their parents do not have adequate/relevant information on sexual and reproductive health. They asserted that the sexuality knowledge of their parents were archaic/out-dated. A male adolescent reported that:

> I don’t discuss sexual issues with my parents especially my mom because they are both ‘old school’. Whatever information they have will be archaic. It’s either I discuss with peers/friends or use the internet

###### Male Adolescent (16-20 years)

One of the female adolescents affirmed same:

> I don’t think my parents are well equipped to give me adequate knowledge on SRH issues. They are archaic. They do not just have enough knowledge. At least I receive up-to-date information in school from my teachers, peers and sometimes from the internet.

###### Female Adolescent (11-15 years)

Adolescents’ reported that their parents were not well equipped to give updated information on sexual and reproductive health issues hence they shy away from such discussions with them. They also narrated meeting their peers and the internet for information.

###### Parental Belief that discussion will initiate sexual intercourse

Some of the adolescents reported that they have not received sex education from their parents because their parents believed it would expose them to sexual exploitation. One of the adolescent avowed:

> Whatever information I got first about sex was from the school. My parents have never discussed sexual issues with me because they probably feel that I would want to venture into sex.

###### Male Adolescent (11-15 years)

In affirmation, another male adolescent narrated that:

> The first time I asked my dad about sex, he was very angry and asked me if I had already started having sex or want to have sex. He also assumed that I had already started having sex when I first asked him about condoms. He almost beat me up because of that one simple question. I was only between 11-15 years old at the time and as a result I vowed never to discuss sexual issues with my dad and he doesn’t discuss with me either. I learn from peers and the internet.

###### Male Adolescent (16-20 years)

Parental assumptions that sexual education leads to initiation to sexual intercourse was reported by the adolescents as barriers to sexual-risk communications.

##### B. Individual factors

###### Lack of trust

All female adolescents agreed that lack of trust between parents and the adolescents hindered effective sexual and reproductive health discussion. Some of their words are captured below:

A female respondent related that:

> I don’t trust my mom and my mom just doesn’t trust me to have these kinds of discussions with me. She feels having these discussions is a way of encouraging me to venture into sex which isn’t true at all.

###### Female Adolescent (16 −20 years)

Another female adolescent affirmed that:

> My parents do not trust my friends because they feel they will influence me negatively and as a result they do not trust me either. I have been warned against having a boyfriend or having sex. If I dare broach the subject of sex to them, then I’m looking for trouble

###### Female Adolescent (11-15 years)

Lack of trust by their parents and lack of trust parents had on their friends was reported by the adolescents as hindrances to sexual risk communication by their parents.

###### Feeling of discomfort with sexual discussion

Some of the adolescents reported that they felt uncomfortable having sexual discussion with their parents. They also noted that their parents felt very uncomfortable too with such discussions. A female adolescent reported that:

> Whenever I ask my Mom about sexual issues, she gets so uncomfortable with the discussion that after a while I stopped discussing with her but I discuss with my friends in school.

###### Female Adolescent (16-20 years)

A male adolescent asserted same:

> My parents rarely discuss sexual issues with me because they are always so uncomfortable with the discussion which also makes me uncomfortable too. I think we all noticed this and stopped its discussion completely. They don’t discuss sexual matters with me anymore.

###### Male Adolescent (16-20 years)

Sexual risk communication discomfort was reported by the adolescents as barriers to sexual education. As a result of this discomfort they sourced sexual and reproductive information from their friends.

###### Fear to initiate sexual with discussion

Some of the adolescents reported being too scared to initiate discussion with their parents. A female adolescent averred that:

> I discuss more of SRH issues like physical development, menstruation with my mom because we are of the same gender. I dare not mention sex or any related topic to her because she will beat me. She will think that I have a boyfriend and want to experiment with him.

###### Female Adolescent (16-20 years)

A male adolescent corroborates same:

> I would love to discuss sexual issues with my parents but I am too scared of what they will think or feel about me.

###### Male Adolescent (16-20 years)

Some of adolescents reported being selective on sexual and reproductive health issues to discuss with their parents as a result of fear associated with sexual discussions.

##### C. Religious/Cultural factors

Some of the respondents revealed that their parents do not discuss sex with them because it is against their religion, cultural or traditional affiliations.

One of the female adolescent narrated that:

> ….my parents are just too religious. Such discussion is a forbidden at home. If I even ask them by mistake, they will skin me alive because they will assume that I want to start experimenting. I’m not even allowed to have male friends around me.

###### Female Adolescent (16-20 years)

A male adolescent affirmed that:

> My parents do not discuss sexual issues with me or my siblings because it is against our religion. Our Church forbids such discussions in the home. Whatever I know about sexual issues, I got from my friends in school.

###### Male Adolescent (16-20 years)

Cultural factors were highlighted as a hindrance to sexuality education. A female adolescent avowed that:

> ….whenever I ask my mom about issues relating to sex, she always tells me it is against our culture to discuss such issues with children.”

###### Female Adolescent (11-15 years)

Adolescents reported that their parent considers sexual education as a taboo as a result of their religious and cultural affiliations.

###### Perceived Barriers to Parent-adolescent Sexual-risk Communication - Parents’ Perspective

When it comes to the necessity of discussing sexual and reproductive health issues with adolescents, parents had a whole lot to say on this. Some of the parents were in the opinion that children may make mistakes if sexuality education isn’t discussed with them and were in agreement that these discussions are needed while a few felt that sexual education isn’t necessary. They perceived that these discussion will give children ideas on sex as children listen more to their parents than they do other persons, they also perceived that discussion will initiate sexual experimentation/early sexual debut.

In agreement on the important of sexuality education to adolescents, one of the parents reported that:

> …discussion with children is very necessary because parents underplay the influence they have on their children’s decisions.

###### Female Parent of Adolescent, Teacher

In corroboration, another parent narrated negative consequences of non-sexuality communication:

> I think parents should discuss with their children. My neighbour’s daughter got pregnant as a teenager and that brought shame to the family. She had to quit going to school at a point as a result of this. Maybe, if her parents had discussed these things with her, she won’t have made such a mistake. As a result of this, I had to start discussing sex with my children. Children should be armed with the knowledge to make wise decisions regarding their sexual life

###### Male Parent of Adolescent, Civil Servant

In disagreement, one of the parents opined that:

> No, discussion on sex with my children will give them ideas to experiment. I don’t discuss it with my children.

###### Male Parent of Adolescent, Trader

A female parent averred that:

> No, I don’t discuss with my children though because I am not comfortable having such discussion with them

#### Female Parent of Adolescent, Petty Trader

Parents narrated their perceived barriers to sexuality communication with the adolescents. These barriers were grouped into three factors namely; Individual factors (shame initiating sexuality discussion and gender barriers, barely enough time for discussion, level of education of parents), adolescent factors (fear that children will want to experiment sexually, feeling that children are too young to be educated on SRH issues), and tradition/religious beliefs.

##### A. Individual factors

###### Shame initiating sexuality discussion and gender barriers

Parents revealed that they sometimes experience a feeling of shame while discussing SRH issues with their children. A female parent reported:

> I have never discussed things like puberty with its associated changes in males and females with my children because i feel so shamed discussing these issues with them it is an abomination in my culture too. My mom never discussed with me so, i never do with my children.”

###### Female Parent of Adolescent, Petty Trader

Gender barriers were one of the factors that hinder parents from effectively discussing sexual and reproductive health issues with their adolescents as revealed in the study. Parents fail to discuss with their children of the opposite sex on sexuality education such as condom use, puberty and physical development. A female parent averred that:

> I have 2 sons who I can’t communicate on sex, condom use or things like ‘wet dreams’ because it is shameful for me. I leave this discussion for my husband to do.

###### Female Parent of Adolescent, Teacher

A male parent had this to say:

> …..I really feel is awkward discussing with my daughters especially on puberty and menstruation. It is the job of their mother and their teachers

###### Male Parent of Adolescent, Civil Servant

Shame initiating sexuality discussions and gender barriers were one of the factors that hinder parents from effectively discussing sexual and reproductive health issues with their adolescents.

###### Barely enough time for discussion

Another barrier that was highlighted by parents was the issue of time. Some of the parents do not discuss with their children because they are just too busy with work and so the discussion is usually left to the other spouse.

A male parent gave this as a reason:

> …..my kind of job doesn’t give me enough time to discuss with my children. I’m usually really tired when I get home and I work most weekends. I only make out time to discuss really important stuff with them.

###### Male Parent of Adolescent, Civil servant

Another corroborated same:

> I don’t have that kind of time. My husband is late so, the whole responsibility of catering for the family falls on me. It’s not easy to be the only parent so, I’m so busy. My child is also a male so; discussing SRH issues with him is not something I’m comfortable with.

###### Female Parent of Adolescent, Cleaner

Parents reported not having enough time for sexuality education and abandoning the sex education for their spouse who may barely have the time to initiate such discussions.

###### Level of education of parents

Another striking barrier is the level of education of parents. Parents who had a higher level of education or work in a corporate establishment discuss more about SRH especially sexual issues than others who had a lower level of education. A male parent with secondary school education reported that:

> I have never discussed sex education with my children because i feel they shouldn’t know about these kinds of things. What they don’t know will not ‘spoil’ them.

###### Male Parent of Adolescent, Trader

A female parent also with secondary education narrated the disadvantage of sex education:

> I have never discussed sex education with my son. I don’t think it’s important that he knows about sex. He may want to experiment if i start discussing with him.

###### Female Parent of Adolescent, Cleaner

Some of the parents who had secondary education never had discussion of SRH issues with their adolescents because of fear of negative impact from the discussion.

##### B. Adolescent factors

###### Fear that children will want to experiment sexually

Some of the parents expressed fear as the reason why they do not discuss SRH issues with their adolescents.. A parent reported that:

> Discussing issues like condom use, sex, STIs/STDs will make my sons feel that they can venture into sex even before they come of age. This is why I don’t discuss these with them.

###### Female Parent of Adolescent, Teacher

Another parent had this to say:

> Discussing things like pregnancy and prevention, relationships etc. will make my daughter feel that it’s okay to have sex in her relationships with the opposite sex probably. Children of this age just want to experiment and besides, she is also too young to understand.

###### Female Parent of Adolescent, Trader

Parents feared that discussion about pregnancy, how to practice safe sex, condom use and sexually transmitted infections will push or encourage their children into engaging in sexual activities

###### Feeling that children are too young to be educated on SRH issues

Most parents do not discuss SRH issues with their children because they feel that their children are too young to be taught SRH issues like sex, pregnancy, condom use, STIs/STDs.

A female parent expresses:

> My daughter is too young for this sort of discussion and I don’t really feel comfortable discussing these kinds of things with her. How can I start educating my less than 15 year old daughter about pregnancy, sex or even STIs? I can’t discuss these with her at such a young age.

###### Female Parent of Adolescent, Petty Trader

Another female parent had this to say:

> I have never discussed sex with my children because they shouldn’t know about sex at their age. They are too young for that

###### Female Parent of Adolescent, Trader

Adolescents especially at their early adolescence were perceived to be too young to receive sexual education by their parents.

##### C. Tradition/Religion

Tradition and religion were also seen as barriers to effective parent-adolescent sexual risk communication. A male parent inferred that:

> For me it’s against my tradition to discuss these things with my children especially my son who is too young. He’s less than 15 years. I feel he should know stuff like these when he’s officially an adult.

###### Male Parent of Adolescent, Trader

Another parent extrapolated that:

> I have never discussed sexual issues with my children because the church frowns at it. I only discuss things like menstruation with my daughter because she needs to know about it and physical development for all my children because they all need to know.

###### Female Parent of Adolescent, Petty Trader

Some of the respondents reported that they do not discuss sexual issues with their children because tradition and region doesn’t allow it. For some of the female parents they only discussed menstruation and physical development with their daughters.

###### Facilitators of sexual and reproductive health discussion with adolescents

Parents were asked what would make it easier for them to discuss SRH issues with their adolescents. The facilitators as listed by the parents were grouped into three; adolescent factors (attaining maturity, closeness with adolescents, and initiation of discussion by adolescence), eliminating barriers (prior discussion by their own parents, permission by the traditional and religious organizations) and parents’ inclusion in sex education (integration of parents on SRH discussions in school).

##### A. Adolescent factors

###### Maturity of the Adolescent and Closeness with children

Some of the parents believed that attaining maturity by the adolescents would necessitate easier adolescent sexual and reproductive health discussions. A parent stated that:

> My children being old enough will make it easier to discuss with them. They are too young right now and may not understand the importance of discussion.

###### Male Parent of Adolescent, Trader

Another parent asserted:

> Discussing sexual issues with my children will probably be easier if they initiated the conversation themselves.

###### Male Parent of Adolescent, Teacher

Another parent believed that parent-adolescent relationship and closeness would facilitate easier adolescent sexuality communication. A male parent agreed that:

> I’m not very close with my children so, they barely discuss this with me or i with them. Closeness with them would have made discussion really easy.

###### Male Parent of Adolescent, Civil servant

Adolescent attaining maturity, adolescents initiating sexuality conversation, closeness with adolescents were some of the adolescent facilitators to initiating sexual-risk communication.

##### B. Eliminating sex education barriers

###### Receiving prior communication from parents

A prior sexuality discussion with parents was suggested to facilitate easier sexual and reproductive health discussions with their children.

A female parent had this to say:

> If my own parents had discussed this with me as an adolescent, maybe discussing with my own children won’t be so difficult or uncomfortable.

###### Female Parent of Adolescent, Teacher

Eliminating barriers in parental sexual-risk communication was reported to facilitate easier communication with adolescents.

###### Permission by traditional and religious organizations

Some of the parents reported that cultural and religious factors impeded free sexuality conservation with their wards.

A parent averred that:

> If tradition and my church don’t forbid it, I would probably discuss with my children on these matters but as it stands now, my hands are tied.

###### Female Parent of Adolescent, Petty Trader

Parents recommend that traditional and religious bodies should encourage sexuality communications to the adolescents.

##### C. Parents inclusion in sex education

###### Integration of parents on SRH discussions in school

Parents mentioned that being included in SRH programmes in their children’s school would go a long way in reminding parents of the importance of discussing SRH issues with their children.

A male parent related that:

> I am a teacher and in the course of my work, I discovered that most parents do not discuss sexual matters with their children unless something bad happens. I think schools should include parents in sex education especially during PTA meetings. Parents can then be encouraged to discuss sexual matters with their children.

###### Male Parent of Adolescent, Teacher

A female parent affirmed that:

> Even though I don’t discuss sexual matters with my children, i feel that if schools invite parents over and discuss this with them, they will be more open to also discuss with their children.

###### Female Parent of Adolescent, Trader

Parents reported that integrating sexual and reproductive health discussions in school using avenues like the Parents Teachers Association (PTA) would facilitate sexuality educations with adolescents at home.

## DISCUSSION

Sex education plays a vital role in the sexual and reproductive health of adolescents; however, parents find it difficult to initiate sexual-risk communications. Adolescents’ involvement in risky sexual behaviours places them at risk of serious unhealthy consequences not only for them but for their parents and the entire community at large. This study was conducted in Rivers State, Nigeria to determine the proportion of adolescents that received sex education from their parents, explore perceived parent-adolescent barriers to sexual-risk communication and factors that may facilitate parent-adolescent sexual-risk communication.

This study revealed that a quarter (21.0%) of the parents had never discussed sex education with their children; however, one-third (32.5%) of the adolescents reported being more comfortable discussing sexual-related issues with their mothers when communication occurs. A higher proportion was found in a study conducted in the USA from 2011–2015 were 57% of adolescent males and 43% of adolescent females did not receive information about sexual birth control or sex education before they first had sex. ^25^ A lower proportion was found in a survey done in southern Ethiopia, which revealed only a low proportion of adolescents communicated with their parents on SRH issues. ^12^ In corroboration, a study carried out in Nigeria revealed that the prevalence of communication between parents and children is low on SRH topics such as HIV/AIDS, family planning, and contraception, at 37.4%, 32.5%, and 9% respectively.^26^ Similarly, another study conducted in Ebonyi State, Nigeria also revealed that the majority of adolescents never discussed anything related to sex with their parents or caregivers but discussed it with their friends or peers. ^27^ The similarities in these studies may be due to the barriers associated with parent-adolescent sexuality communications. This low parent-adolescent sexual and reproductive health communication can make adolescents more prone to diverse negative sexual and reproductive health outcomes.

With the low parent-adolescent sexual and reproductive health communications observed in this study, the barriers were explored from both the adolescent and parent’s perspective. Adolescents outlined some of the perceived barriers to effective sexual-risk communication grouped into three broad factors namely, parental factors (parents being too busy, parents being too judgmental, parental poor/inaccurate knowledge on sex education, parental belief that discussion will initiate sex); individual factors (lack of trust, fear/discomfort with discussion) and religious/cultural factors. Focusing on the adolescent perspectives, both the male and female adolescents jointly agreed from their different focus group discussions that their parents were too busy to initiate sexual and reproductive health communications with them. Parents left home most times early in the morning and returned home late at night tired. They barely had time to interact with the adolescents or even engaging in sexual-risk communications. This was in line with a qualitative review in East Africa that showed parents had no time to interact with the adolescents especially the publically employed with tight working schedules, unlike the self-employed parents who had more time to discuss with the adolescents. ^28^ In agreement, another study revealed that adolescents felt disconnected from their parents due to the fact that they are always busy with work or never home resulting in parent-adolescent sexuality barriers. ^29^ This reveals the need for work-life balance to enable parents to spend more time with their children and discuss sexual and reproductive health matters with them. The adolescents also indicated the information they obtained on sex education was gotten from the schools, their peers and from internet sources. This was similar with finding conducted by Krugu et al. ^30^ and Motsomi et al. ^29^ in Ghana and South Africa respectively.

The focus group discussions with the adolescent also revealed female adolescents do not discuss sexual matter with their parents because parents are too judgmental and would feel that they are already engaging in sex or want to initiate sexual intercourse through discussions on sexual matters. A study conducted in South Africa revealed same, it was reported that female adolescents found it difficult to face their parents after such a discussion and they thought their mothers would condemn them and assume they had started sleeping around with boys. ^29^ In the same vein, some of the male adolescents reported that they asked their fathers’ about condom use and its effectiveness in prevention of pregnancy and they received a shocking feedback, they were either beaten or interrogated further in a condensing manner. With these judgmental attitudes by parents, most of the adolescents never initiated such conversations again; they relied on other sources of information. Health education may be useful in correcting this poor knowledge and judgmental attitude of parents.

Most of the adolescents believed that their parents had poor or inaccurate knowledge on sex education. They felt that their parents were either “old school” or have archaic knowledge on sexuality matters hence they relied on other updated sources especially the internet and social media platforms. In agreement, a study conducted in Ibadan reported that parents felt their children thought they were ‘‘old school’’ (traditional in views) and may judge them when they initiate a conversation around SRH issues.^31^ This finding was in corroboration with a study carried out in Ebonyi State, Nigeria which revealed that parents reported that they do not know what to discuss when it comes to sex-related matters. ^32^ Similarly, another study revealed that parent was unsure of right answers to questions on sex which served as a barrier to sexuality communications. ^33^ The similarities in these studies reveal the need for enhanced training in sex education for parents and guardians.

Following the individual factors, adolescents reported they trusted their parents less with sexual-risk communications. Also, most of the female adolescents reported that their parents do not trust them because of the friends they keep. Their parents feared they were already sexually active or that such communication could lead to future experimentation by the adolescents. This was in line with a study that reported barriers to sexual communication; it was noted that sex education was perceived to lead to an early sexual debut and has no positive effect on adolescent behaviour. ^33^ A few reported that the only sexual and reproductive health communication they had with their parents was on physical development and menstrual hygiene. The adolescents further revealed that they were uncomfortable engaging in sexuality discussions with their parents, and some of the adolescents were too scared to initiate such discussions. Similar findings from a study revealed barriers to effective sexual risk communication, including discomfort with discussion and disinterest in adolescents. ^33^ Health education and enhancing parent-child relationships could limit these trust and misconception barriers.

The cultural and religious inclinations of their parents were reported by both genders as stumbling blocks to sex education. Adolescents reported that their parents vehemently refused to engage in sex education with them, even when they tried to initiate the conversation. Culturally, it was perceived as taboo to be discussed, and religiously, it was perceived to lead to sin. This finding was in agreement with a study conducted in Ghana, which revealed that the majority of rural adolescents do not communicate with their parents or caregivers with regard to sexual issues due to cultural and religious norms that prohibit adolescent-parent sexual communication.^34^

Parents also expounded on some problems they encounter while discussing sexual and reproductive health matters with their adolescents. When parents were asked if they agreed to initiate sexuality education for adolescents, varying opinions were received. Most of the parents who were traders were in disagreement while the teachers and civil servants were in agreement on discussing sexuality education with the adolescents. Similarly, a study revealed that parents who were civil servants were more likely to initiate sexuality communication than those in other groups. ^35^ This may be due the exposure that stems from working in an enlightened organization which could increases the knowledge of parents on the necessity of sexuality education. The parents who were in agreement to initiate sexual and reproductive health communications further expressed that a lack of sexuality communication could lead to more deleterious sexual and reproductive outcomes.

Parents narrated their perceived barriers to sexuality communication with the adolescents. These barriers were grouped into three factors, namely: individual factors (shame initiating sexuality discussion and gender barriers, barely enough time for discussion, level of education), adolescent factors (fear that children will want to experiment sexually, feeling that children are too young to be educated on SRH issues), and tradition/religious beliefs. Parents expressed being shy and having discomfort initiating sexuality and reproductive health discussions. A study conducted in South Africa revealed that most parents expressed discomfort and experienced embarrassment discussing sexual issues with their children, and they fear being misconstrued as wanting to engage in sexual activity with them. ^29^ However, one of the female parents revealed she never received such discussions from her mother and would never initiate such discussions due to shame and embarrassments that emanates from such conversations. In agreement, a study revealed parental barriers as parental discomfort discussing sex. Health education could be helpful to bridge these communication barriers. ^36^

Gender barriers were outlined as one of the hindrances parents face to effective discussion on sexual and reproductive health issues with their adolescents. Parents find it difficult to discuss with their children of the opposite gender sexual and reproductive health topics such as condom use, puberty, and physical development. This poses a problem as parents of both sexes tend to communicate with children of the same gender; fathers would rather discuss with their sons, while mothers do the same for their daughters. In corroboration, a qualitative review conducted in East Africa reported gender barriers in that mothers and fathers of adolescents in this region discuss more with adolescents of the same sex because both parents feel shy and hence find it difficult to openly talk to their children. ^28^This poses a need to address gender norms and stereotypes as it creates a stumbling block to parent-child sexual-risk communications.

Another barrier that was highlighted by parents was the issue of time. Some of the parents do not discuss this with their children because they are just too busy with work. In most cases, they assume the other partner would engage in such a discussion with the adolescent. Parents who had attained a higher level of education or worked in a corporate establishment discussed more sexual and reproductive health issues with their adolescents than others who had a lower level of education. This corroborates findings which revealed that parents with a tertiary level of education were more likely to initiate sexual and reproductive health communication than those with a primary level of education. ^34^ A qualitative review in East Africa showed that educated parents had the patience to talk verbally and face-to-face with their adolescents as compared to parents with less or without education. ^28^ More health education and enlightenment campaigns would be useful to break these barriers.

Most of the parents expressed fear in initiating sexuality education with the adolescents. They feared that discussions about pregnancy, how to practice safe sex, condom use, and so on would push or encourage their children to engage in sexual activities. This was in line with a study that revealed that parents felt that engaging in discussions about sexuality and reproductive health issues would encourage their children to indulge in sex. ^29^ Most parents do not discuss sexual and reproductive health issues with their children because they feel that their children are too young to be taught about issues like sex, pregnancy, condom use, and STIs, they also believe they are too young to comprehend. They believed that the knowledge would come naturally to them as they grow into adulthood. This was similar to a finding from South Africa, which reported that most parents cited discussing issues pertaining to puberty, condom use, sexually transmitted infections, and contraceptives as inappropriate because their children are still too young. ^29^ Another study affirmed that a child being too young served as a hindrance to parent-adolescent sexuality communications. ^32^ Nevertheless, Nundwe ^22^ indicated that effective interaction on sexual and reproductive health is likely to reduce adolescent risk-taking sexual behaviours when parents talk with adolescents on sexuality issues.

Cultural and religious beliefs were also seen as inhibiters to effective parent-adolescent sexual risk communication. Some of the parents reported that they do not discuss sexual and reproductive health matters with their children because their traditional and religious belief frowns at such discussions. In the same vein, Malaclane and Beckmeyer ^35^ revealed parental barriers to include: racial/ethnic and religious factors. This was similar to studies conducted in South Africa and East Africa, which showed that traditional norms, culture and religious beliefs prohibited parents to speak on issues regarding sexuality to adolescents as it’s seen as an abomination and a taboo hence they shy away. ^28, 29^ This may possibly indicate parents’ incapability to disentangle from the traditional and religious norms around sexuality which is believed that such discussion could lead adolescents into sexual explorations. This further emphasizes the need to change the culture of silence around sexuality and educate adolescents appropriately on sexual and reproductive health issues.

Facilitators for easier parent-adolescent sexual and reproductive health discussion were also explored. Parents suggested that prior discussion on sexuality communications by their own parents, permission by the church, removing traditional barriers, initiation of discussion by adolescence, adolescents attaining maturity, having a closer relationship with the adolescents and integration of parents on sexual and reproductive health discussions in school would help facilitate sexuality discussions. Adolescence is a crucial time for socialization and one important aspect of socialization is communication about sexual and reproductive health matters. ^36^ The earliest and critical influencers on the sexual development of children are parents which make them most suitable source of provision of sex education to their wards due to extensive involvement in their lives. ^24^ Parents have the ability to guide their children’s sexual and reproductive health development, and also encourage them to practice safe sexual behaviour. ^13^ In facilitating parent-adolescent sexual-risk communications, parents must learn to interact socially with the adolescents.

## Conclusion

The current study revealed that a quarter of the parents had never discussed sex education with their children, and one-third of the adolescents reported being more comfortable discussing sexual-related issues with their mother. It was shown that more than half received sexual education from school; others were from home, the internet, or social media. From the adolescent perspective, adolescent perceived barriers to sexual risk communication include adolescents being scared or uncomfortable discussing sex with their parents, parents being too busy, parents being too judgmental, a lack of trust by parents, belief by parents that sexuality discussions would lead to early sexual initiation, parents having poor or inaccurate knowledge about sex and cultural or religious barriers. Parents perceived barriers to adolescent sexual risk communication as gender differences or barriers, shame initiating sexuality discussion, fear that children will want to experiment sexually, feeling that children are too young to be educated on SRH issues, inadequate knowledge of SRH topics, lack of time for discussion, level of parental education, and traditional or religious beliefs. Understanding these barriers would help in improving parent-adolescent sexuality communications in Rivers State and in Nigeria at large.

This study had its limitations, due to the sensitivity of the topic; some of the adolescents were shy during interview sessions. To curb some of these limitations, the adolescents were assured of the confidentiality of the information divulged. The parents that engaged in the in-depth interview gave the same responses; thus, we achieved saturation on time, hence the small sample size. Finally, this study was conducted among in-school adolescents and may not be generalizable for out-of-school adolescents. Despite these limitations, the study contributed a lot to the literature by expounding on parent-adolescent barriers to sexual risk communication in Nigeria.

We recommend that adolescents should also be open to having an open and honest discussion about sex with their parents so as to minimize risky sexual behaviour. Parents should be trained to start sexuality education especially at early adolescence. The government should organize training for parents to enable them overcome the barriers to discussing sexual and reproductive health issues with their children and also on how to properly communicate with their children of the opposite sex. Programs to address socio-cultural norms and religious beliefs that act as stumbling blocks to communication could be useful. The Ministry of Health and Education should also organize school health programmes to coach parents and adolescents on how best to overcome the barriers to parent-adolescent sexuality communication.

## Data Availability

The data that support the findings of this study are not openly available for ethical reasons (underage human participants), and are available from the corresponding author upon reasonable request.

